# Immunogenicity and safety of COVID-19 vaccine in lung cancer patients receiving anticancer treatment: A prospective multicenter cohort study

**DOI:** 10.1101/2022.06.23.22276536

**Authors:** Kei Nakashima, Masayuki Ishida, Hiroyuki Matsui, Chihiro Yoshida, Tatsuya Nagai, Minoru Shiraga, Hiroshi Nakaoka, Yoshihito Otsuka, Yu Nakagama, Natsuko Kaku, Yuko Nitahara, Yasutoshi Kido, Yoshio Hirota

**Affiliations:** Department of Pulmonology, Kameda Medical Center, 929 Higashi-cho, Kamogawa, Chiba 296-8602, Japan; Department of Pulmonology, Chikamori Hospital, 1-16 Okawasuji, Kochi, 780-8522, Japan; Clinical Research Support Office, Kameda Medical Center, 929 Higashi-cho, Kamogawa, Chiba 296-8602, Japan; Department of Laboratory medicine, Kameda Medical Center, 929 Higashi-cho, Kamogawa, Chiba 296-8602, Japan; Department of Parasitology and Research Center for Infectious Disease Sciences, Graduate School of Medicine, Osaka City University, 1-4-3 Asahi-machi, Abeno-ku, Osaka 545-8585, Japan; Clinical Epidemiology Research Center, SOUSEIKAI Medical Group (Medical Co. LTA), 3-5-1 Kashii-Teriha, Higashi-ku, Fukuoka, 813-0017 Japan

**Keywords:** COVID-19 vaccine, Lung cancer, Anticancer treatment, Immunogenicity, Vaccine safety

## Abstract

**Introduction:** This study assessed the immunogenicity and safety of BNT162b2 mRNA vaccine in lung cancer patients receiving anticancer treatment using two immunoassays. **Methods:** We enrolled lung cancer patients receiving anticancer treatment and non-cancer patients with chronic diseases; all participants were fully vaccinated with the BNT162b2 vaccine. Blood samples were collected before the first and second vaccinations and 4 ± 1 weeks after the second vaccination. Anti-acute respiratory syndrome coronavirus-2 (SARS-CoV-2) spike protein S1 subunit receptor-binding domain antibody titers were measured using the Architect SARS-CoV-2 IgG II Quant (Abbott Laboratory) and Elecsys Anti-SARS-CoV-2 S (Roche Diagnostics).

**Results:** Fifty-five lung cancer patients and 38 non-cancer patients were included in the immunogenicity analysis. Lung cancer patients showed significant increase in the geometric mean antibody titer, which was significantly lower than that in the non-cancer patients after the first (30 vs. 121 AU/mL, p<0.001 on Architect; 4.0 vs 1.2 U/mL, p<0.001, on Elecsys) and second vaccinations (1632 vs. 3472 AU/mL, p=0.005, on Architect; 213 vs 573 A/mL, p=0.002, on Elecsys). The adjusted odds ratio (OR) for seroprotection was significantly lower in the lung cancer patients. Analysis of the anticancer treatment types showed that the adjusted OR for seroprotection was significantly lower in lung cancer patients receiving cytotoxic agents. Lung cancer patients showed no increase in the number of adverse reactions.

**Conclusions:** BNT162b2 vaccination in lung cancer patients undergoing anticancer treatment significantly increased antibody titers and showed acceptable safety. However, the immunogenicity in these patients could be inadequate compared with that in non-cancer patients.

## Introduction

Coronavirus disease (COVID-19), which originated in China in December 2019, spread globally with unprecedented impact.^1^ The COVID-19 pandemic has affected cancer patient management in various ways.^2^ Considering the poor outcome of COVID-19, cancer patients are particularly at risk from the disease.^2^ Most cancer patients are elderly people, who tend to have other underlying diseases and are often immunosuppressed owing to anticancer treatment.^2^ This makes them vulnerable to infection and increases their risk of developing serious complications from the virus, which in turn could lead to hospitalization, admission to intensive care units, and/or death.^3-7^ Additionally, further anticancer treatment has been postponed because of the need to prioritize treatment for COVID-19.^2^ Therefore, prevention of COVID-19 in cancer patients is crucial.

Vaccines are the primary means to prevent COVID-19; moreover, several formulations have been launched since November 2020.^8-10^ Cancer patients are being prioritized for COVID-19 vaccinations globally, taking into consideration their poor clinical outcomes when infected.^11^ In a recent study, the antibody titers in patients with solid cancers were reduced compared with those in healthy controls; however, an adequate antibody response was achieved after two doses of vaccination.^12^ Moreover, the safety profile of the COVID-19 vaccine in patients with solid cancer was found to be acceptable.^13^ However, studies on the immunogenicity and safety of the COVID-19 vaccine specific to lung cancer patients undergoing chemotherapy are insufficient.^14^ Additionally, variations in immunogenicity depending on the type of anticancer drug administered to patients are unclear. Finally, the differences between anti-severe respiratory syndrome coronavirus-2 (SARS-CoV-2) receptor-binding domain spike protein IgG (anti-RBD) titers as measured by two different immunoassays in lung cancer patients are unknown.

Thus, in this prospective multicenter cohort study, we aimed to evaluate the immunogenicity and safety of the BNT162b2 vaccine in lung cancer patients undergoing anticancer treatment using two different immunoassays to measure anti-RBD titers.

## Materials and Methods

### Study participants

Lung cancer patients receiving anticancer treatment and non-cancer patients with chronic diseases at the pulmonology departments of two tertiary hospitals in Japan (Kameda Medical Center, Chiba and Chikamori Hospital, Kochi) were invited to participate in the study from May 2020 to September 2021. The inclusion criteria were patients fulfilling all of the following at the time of enrollment as follows: 1) lung cancer patients currently undergoing anticancer treatment aged ≥ 50 years (lung cancer group), or non-cancer patients with chronic diseases aged ≥ 50 years (non-cancer group); 2) patients who voluntarily received the COVID-19 vaccine; and 3) patients who provided informed written consent to participate in the study. The exclusion criteria were patients with any of the following at the time of enrollment: 1) patients with contraindications to vaccination; 2) those previously vaccinated with a COVID-19 vaccine (including participation in clinical trials or clinical studies); 3) those who had been receiving systemic steroids or immunosuppressive drugs (except for administration as an antiemetic for anticancer drugs); 4) those with autoimmune diseases under active treatment; 5) those who experienced an acute illness requiring antimicrobial agents or steroids within the past month, at the time of enrollment; 6) those with fever or acute severe diseases at the timing of vaccination; and 7) those judged by the principal investigator/sub-investigator to be unsuitable as research subjects. The study protocol was approved by the Hakata Clinic Institutional Review Board (no O-50) and was performed in accordance with the Declaration of Helsinki. This study was registered with the Japan Registry of Clinical Trials, trial number: jRCT1071210024.

During registration, the study participants were asked to fill out a self-administered questionnaire comprising the following information: date of birth, sex, age, height, weight, and allergy history. Additionally, the attending physicians gathered the following information from the participants on the standardized questionnaire: sex, age, underlying diseases, and Eastern Cooperative Oncology Group performance status. The attending physician gathered the following information for lung cancer patients: the histological type of the lung cancer and details of anticancer treatment.

### COVID-19 vaccination

All study participants were administered the BNT162b2 mRNA COVID-19 vaccine (COMIRNATY® intramuscular injection, Pfizer, New York, NY, USA). In Japan, mass vaccination was undertaken for BNT162b2 vaccines, which were stored and prepared according to the package insert. Each person received two vaccination doses, three to four weeks apart.

### Measurement of antibody titers

Blood samples for pre-vaccination antibody titer measurements were collected from study participants within 14 days before the first COVID-19 vaccination (S0); those for post-vaccination antibody titer measurements were collected within 7 days before the second vaccination (S1) and 4 ± 1 weeks (21–35 days) after the second vaccination (S2). The collected serum samples were stored at -20 °C in the laboratory of the respective hospitals.

Titers of anti-RBD and SARS-CoV-2 nucleocapsid protein (anti-N) were measured from the collected blood samples using the following two assays: Architect SARS-CoV-2 IgG II Quant (Abbott Laboratories, Illinois, USA) and Elecsys Anti-SARS-CoV-2 (Roche Diagnostics, Basel, Switzerland).^15, 16^ For the Architect assay, the quantitative range was 0–40,000 (AU/mL) in this study, and the cutoff value for a positive anti-RBD and anti-N test result was ≥ 50 (AU/mL) and 1.4, respectively ^12, 15^. For Elecsys, the quantitative range was 0.4–2,500 (U/mL), and the cutoff value for a positive anti-RBD and anti-N test result was ≥ 0.8 (U/mL) and ≥ 1.0, respectively ^16^

### Safety

Ocular and respiratory symptoms within 48 h after the first and second vaccinations, local reactions at the injection site, and systemic reactions within 7 d after the first and second vaccinations were monitored using case cards completed by the participants.

### Statistical analysis

Study participants positive for anti-N antibodies (i.e., those who acquired COVID-19) in the blood samples before (S0) and after the first (S1) and second (S2) vaccination were excluded. The immunogenicity of the COVID-19 vaccine was measured using the geometric mean antibody titer (GMT) and GMT ratio of the anti-RBD antibody titer. The cutoff point for seropositivity was ≥ 50 (AU/mL) and ≥ 0.8 (U/mL) for the Architect and Elecsys assay, respectively. We set the following two cutoff points for seroprotection according to the previous studies1) ≥ 154 BAU/mL based on the mean protective threshold for the original wild-type strain based on data from six vaccine regimens; 2) ≥ 775 BAU/mL based on the 90% vaccine efficacy threshold against symptomatic COVID-19 reported in a prior clinical trial of mRNA vaccine according to the international standard^17-19^. Although ≥ 775 BAU/mL is considered to be a widely accepted cutoff value, 154 BAU/mL was also adopted from a previous study because the present study included elderly patients with cancer or non-cancer patients, and the geometric mean of the antibody titer of these populations was considered relatively low.^17, 19^ The value of 154 BAU/mL was converted to 1,084 AU/mL (BAU/mL × 7.042) and 775 U/mL (BAU/mL × 0.971), and 775 BAU/mL was converted to 5,458 AU/mL (BAU/mL × 7.042) and 753 U/mL (BAU/mL × 0.971), for the Architect and Elecsys assay, respectively.^18, 20^ During data processing, anti-RBD antibody titer of 0 AU/mL was regarded as 0.1AU/mL for the Architect assay. Anti-RBD antibody titer of < 0.4 AU/mL and >2500 was regarded as 0.4 and 2500 for the Elecsys assay, respectively. Reciprocal antibody titers were analyzed after logarithmic transformation, and the results are presented on the original scale by calculating the antilogarithm. Stratified analyses were performed to examine the effects of the following potential confounders: age at vaccination (<70, 70–74, ≥ 75 years), sex, group (lung cancer group, non-cancer group), and type of anticancer treatment. The significance of the fold rise within a category was assessed using the Wilcoxon signed-rank test; moreover, inter-category comparisons were made using the Wilcoxon rank-sum test of the Kruskal-Wallis test. The Student’s t-test, Fisher’s exact test, Jonckheere-Terpstra test, and Cochran-Armitage test were performed where appropriate. Additionally, the independent effects of potential confounders on antibody induction were assessed using logistic regression analysis. Models were constructed considering seroprotection as the dependent variable and the abovementioned potential confounders as explanatory variables. The ORs and 95% confidence intervals (CIs) were calculated. All tests were two-sided, and all analyses were performed using R version 4.1.2 (R Foundation for Statistical Computing; http://www.r-project.org). Differences were considered significant at p <0.05.

## Results

A total of 55 lung cancer patients and 38 non-cancer patients with chronic diseases were enrolled in this study. One lung cancer patient dropped out, while another was positive for anti-N antibodies on the Architect assay, suggesting a previous infection before the first vaccination. After excluding these two patients, 91 patients completed the three blood collections up to 4 weeks after the second vaccination; they were evaluated for immunogenicity and safety as they were negative for anti-N antibodies in both assays in S0, S1, and S2. All study participants were completely vaccinated with the BNT162b2 COVID-19 vaccine (two doses administered three to four weeks apart).

Table 1 shows a comparison of the characteristics between lung cancer patients and non-cancer patients included in the immunogenicity analysis. There was no difference between the lung cancer patients and non-cancer patients with chronic diseases in terms of their mean age. No differences existed in the proportion of men (53% in lung cancer patients vs. 40% in non-cancer patients), history of allergies (28% vs. 32%), or performance status between lung cancer patients and non-cancer patients. The most frequent underlying diseases were respiratory disease (100% vs. 97%), hypertension (59% vs. 45%), and dyslipidemia (34% vs. 34%) in lung cancer patients and non-cancer patients, respectively. No significant differences in any underlying diseases existed between the two groups. Adenocarcinoma was the most common histological type of lung cancer in 44 patients (83%). The most common anticancer therapy comprised tyrosine kinase inhibitors (TKIs) in 20 patients (38%), followed by immune-checkpoint inhibitors (ICIs) in 11 patients (21%).

**Table 1.**
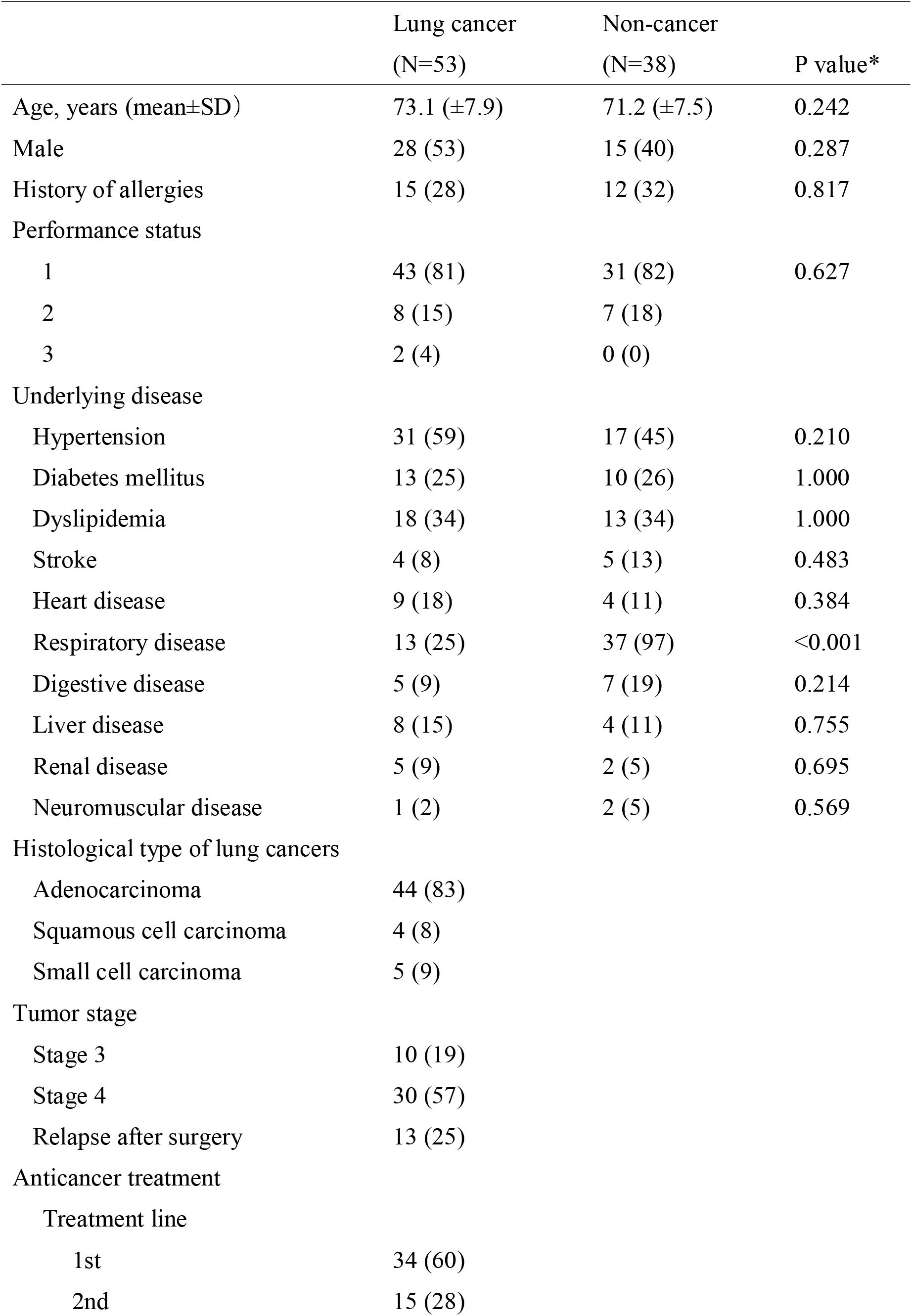
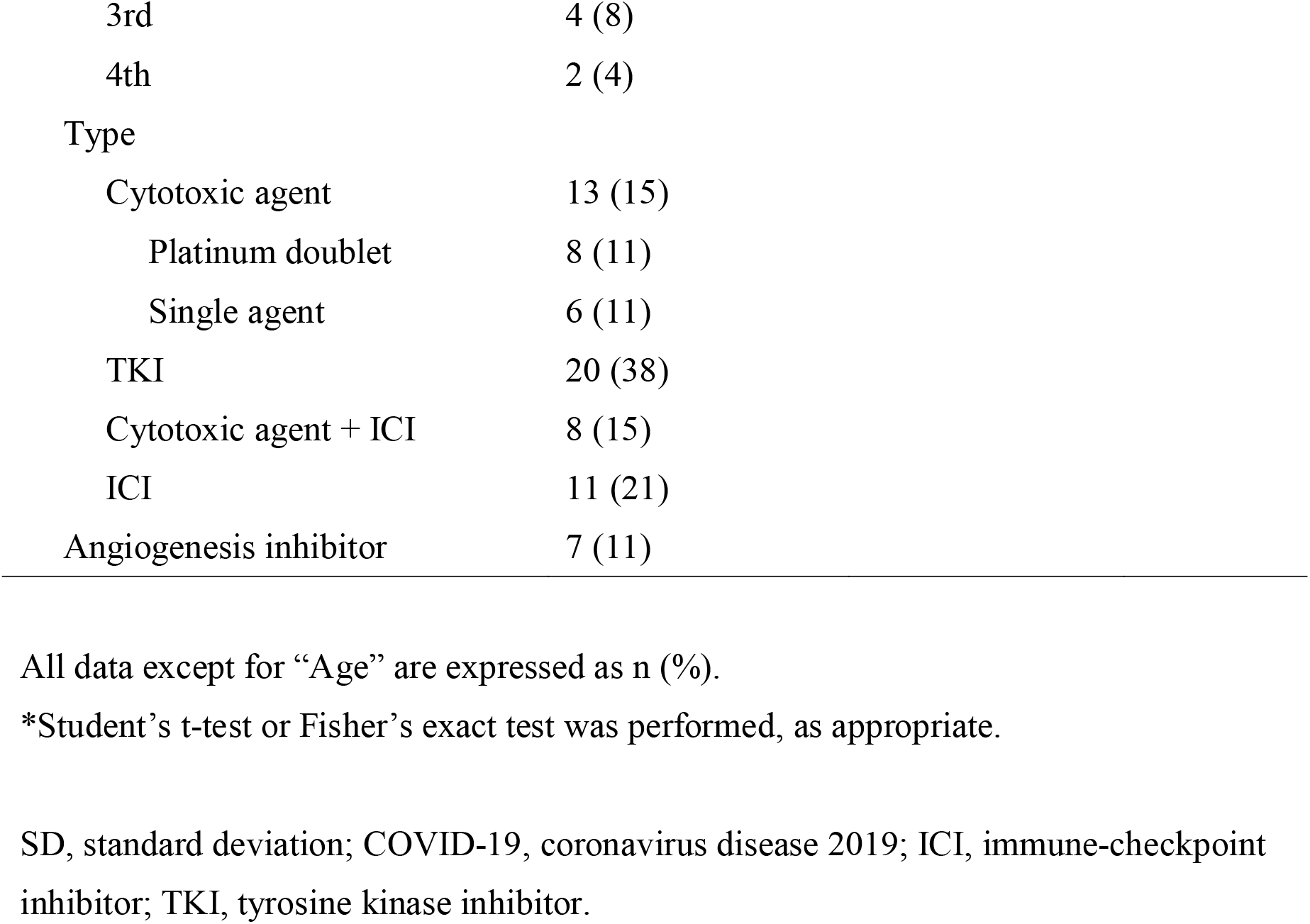
Characteristics of study patients included in the immunogenicity analysis

|  | Lung cancer<br>(N=53) | Non-cancer<br>(N=38) | P value* |
| --- | --- | --- | --- |
| Age, years (mean±SD) | 73.1 (±7.9) | 71.2 (±7.5) | 0.242 |
| Male | 28 (53) | 15 (40) | 0.287 |
| History of allergies | 15 (28) | 12 (32) | 0.817 |
| Performance status |  |  |  |
| 1 | 43 (81) | 31 (82) | 0.627 |
| 2 | 8 (15) | 7 (18) |  |
| 3 | 2 (4) | 0 (0) |  |
| Underlying disease |  |  |  |
| Hypertension | 31 (59) | 17 (45) | 0.210 |
| Diabetes mellitus | 13 (25) | 10 (26) | 1.000 |
| Dyslipidemia | 18 (34) | 13 (34) | 1.000 |
| Stroke | 4 (8) | 5 (13) | 0.483 |
| Heart disease | 9 (18) | 4 (11) | 0.384 |
| Respiratory disease | 13 (25) | 37 (97) | <0.001 |
| Digestive disease | 5 (9) | 7 (19) | 0.214 |
| Liver disease | 8 (15) | 4 (11) | 0.755 |
| Renal disease | 5 (9) | 2 (5) | 0.695 |
| Neuromuscular disease | 1 (2) | 2 (5) | 0.569 |
| Histological type of lung cancers |  |  |  |
| Adenocarcinoma | 44 (83) |  |  |
| Squamous cell carcinoma | 4 (8) |  |  |
| Small cell carcinoma | 5 (9) |  |  |
| Tumor stage |  |  |  |
| Stage 3 | 10 (19) |  |  |
| Stage 4 | 30 (57) |  |  |
| Relapse after surgery | 13 (25) |  |  |
| Anticancer treatment |  |  |  |
| Treatment line |  |  |  |
| 1st | 34 (60) |  |  |
| 2nd | 15 (28) |  |  |

|  |  |
| --- | --- |
| 3rd | 4 (8) |
| 4th | 2 (4) |
| Type |  |
| Cytotoxic agent | 13 (15) |
| Platinum doublet | 8 (11) |
| Single agent | 6 (11) |
| TKI | 20 (38) |
| Cytotoxic agent + ICI | 8 (15) |
| ICI | 11 (21) |
| Angiogenesis inhibitor | 7 (11) |

All data except for “Age” are expressed as n (%).
\*Student’s t-test or Fisher’s exact test was performed, as appropriate.
SD, standard deviation; COVID-19, coronavirus disease 2019; ICI, immune-checkpoint inhibitor; TKI, tyrosine kinase inhibitor.

Table 2 shows the GMT and GMT ratio according to age, gender, group, and the type of anticancer treatment. In all subjects, the GMT of the BNT162b2 vaccine was 54 AU/mL on Architect and 2.0 U/mL on Elecsys after the first vaccination and 2237 AU/mL on Architect and 322 U/mL on Elecsys after the second vaccination, indicating a significant increase. In the stratified analysis considering underlying disease, the GMTs of lung cancer patients were significantly lower than those of the non-cancer patients after the first vaccination (30 vs. 121 AU/mL, p<0.001 on Architect; 4.0 vs 1.2 U/mL, p<0.001, on Elecsys) and second vaccination (1632 vs. 3472 AU/mL, p=0.005, on Architect; 213 vs 573 A/mL, p=0.002, on Elecsys). Figure 1 shows the changed in the anti-RBD antibody titer before vaccination (S0), after first vaccination (S1), and after second vaccination (S2) between non-cancer and lung cancer patients. Regarding types of anticancer treatments, in lung cancer patients, a significant increase in GMT was observed after each of the two doses of the BNT162b2 vaccine for all treatment types on both Arthitect and Elecsys. After the second vaccination, the GMTs varied significantly for the various treatments on both Architect and Elecsys; in particular, lung cancer patients receiving cytotoxic agents showed a low value of 818 AU/mL and 72 U/mL for Architect and Elecsys, respectively. Interestingly, the GMT ratio of S2/S0 was significantly different between the groups and among the types of anticancer treatment on Elecsys, whereas there was no significant difference on Architect; reduced immunogenicity in lung cancer patients was notable in Elecsys assay compared with that in Architect assay. Supplemental Figure 1 shows changes in the anti-RBD antibody titers before vaccination (S0) and after the first (S1) and second (S2) vaccinations among the types of anticancer treatment. Reduction in anti-RBD titers in lung cancer patients receiving cytotoxic agents after the second vaccination (S2) was more pronounced in the results of the Elecsys assay.

**Table 2.**
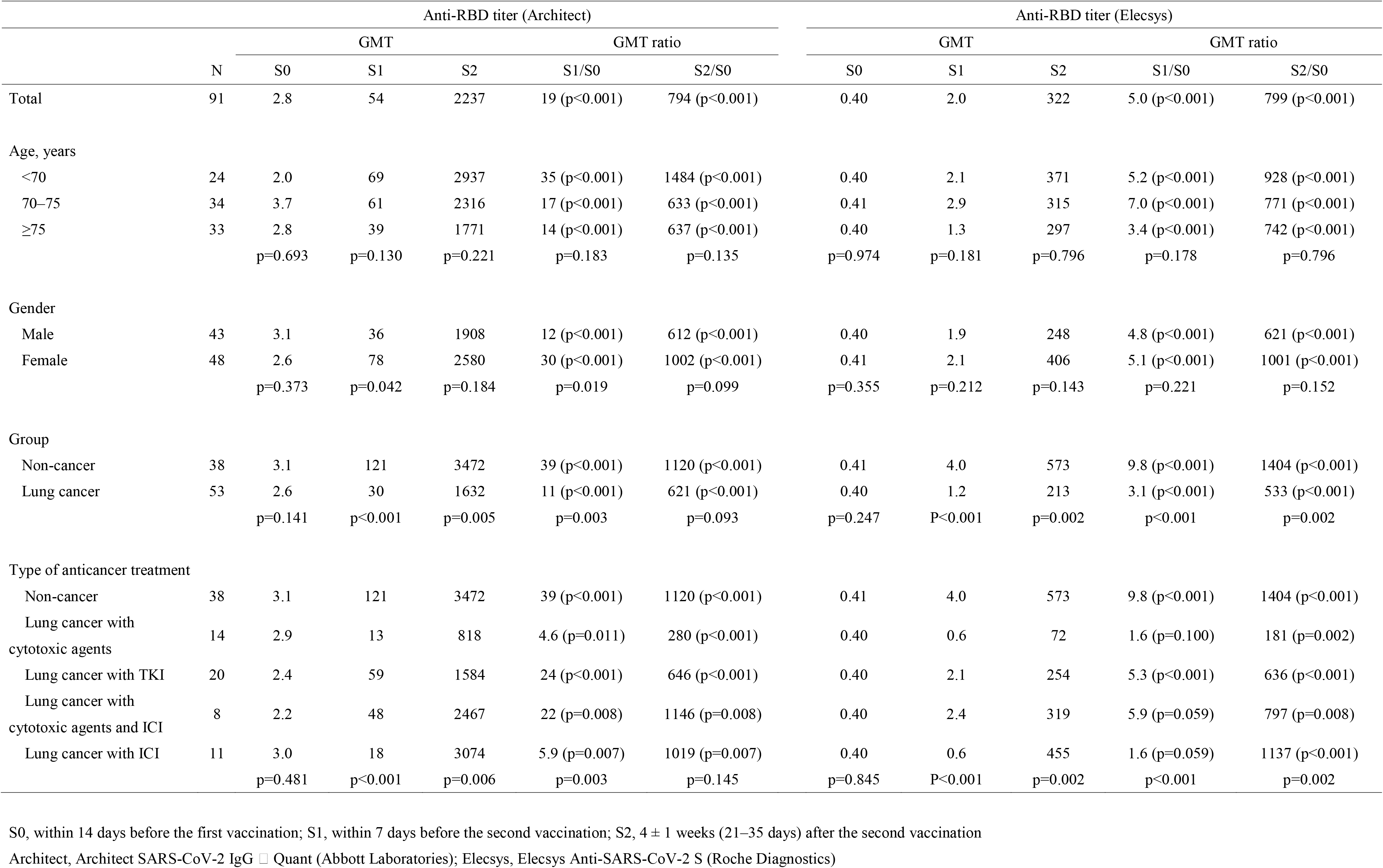
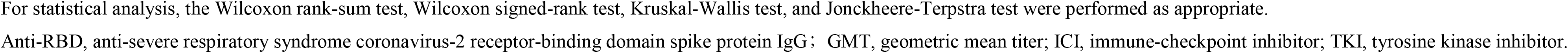
Comparison of the geometric mean titer (GMT) and GMT ratio of the anti-RBD antibodies among study participants stratified by age, gender, group, and the type of anticancer treatment.

**Figure 1.**
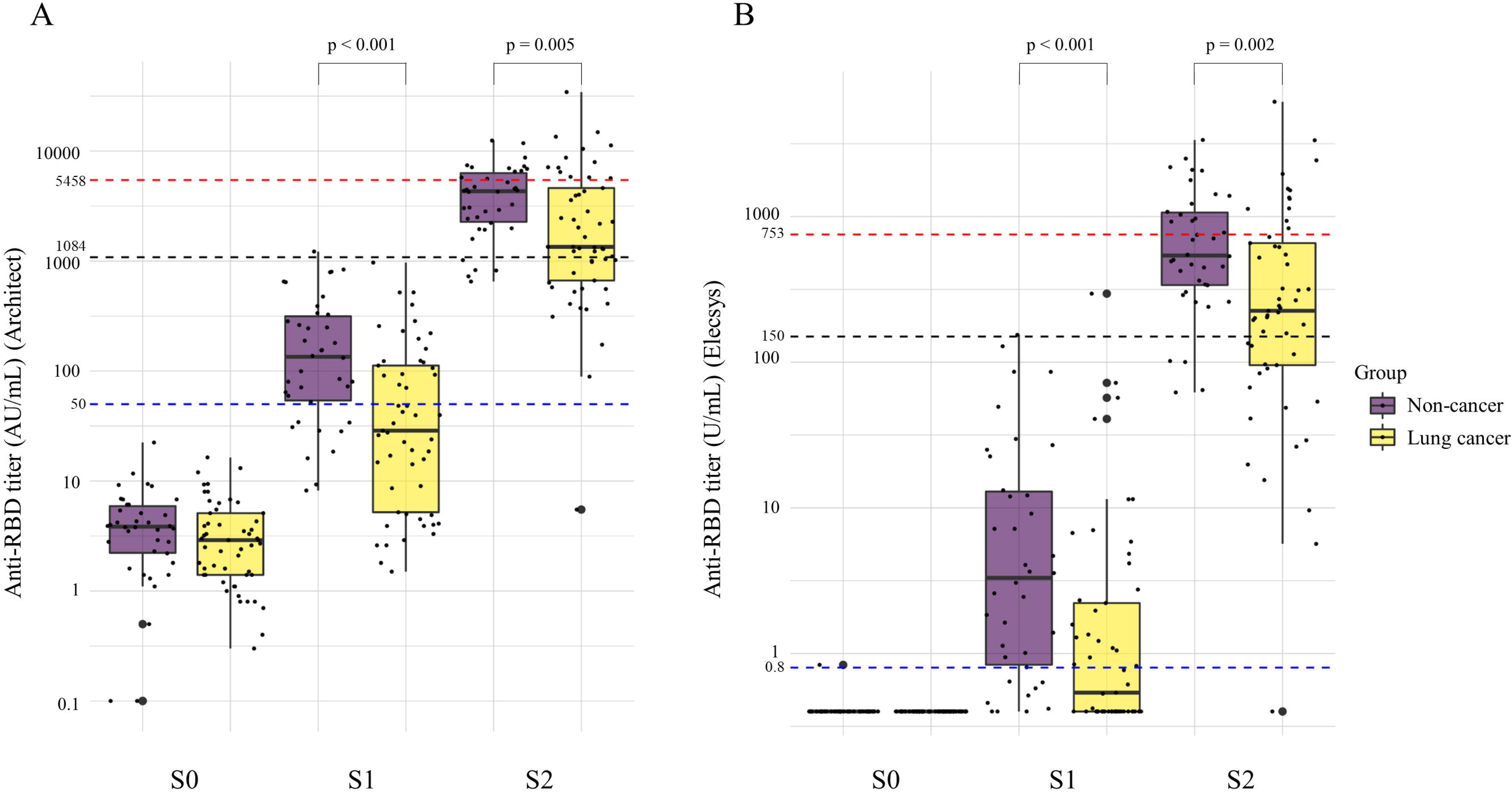
Changes in the anti-RBD antibody titer before vaccination (S0), after first vaccination (S1), and after second vaccination (S2) between non-cancer and lung cancer patients. A Anti-RBD titer on Architect. B Anti-RBD titer on Elecsys. The GMTs of lung cancer patients were significantly lower than those of the non-cancer patients after the first vaccination (30 vs. 121 AU/mL, p<0.001 on Architect; 4.0 vs 1.2 U/mL, p<0.001, on Elecsys) and second vaccination (1632 vs. 3472 AU/mL, p=0.005, on Architect; 213 vs 573 A/mL, p=0.002, on Elecsys). Anti-RBD, anti-severe respiratory syndrome coronavirus-2 receptor-binding domain spike protein IgG S0, within 14 days before the first vaccination; S1, within 7 days before the second vaccination; S2, 4 ± 1 weeks (21–35 days) after the second vaccination Architect, Architect SARS-CoV-2 IgG □ Quant (Abbott Laboratories); Elecsys, Elecsys Anti-SARS-CoV-2 S (Roche Diagnostics)

Table 3 shows the seropositivity and seroprotection according to age, gender, group, and the type of anticancer treatment. Among all subjects, the seropositivity on Architect and Elecsys were 54% and 58% after the first vaccination, and 99% and 99% after the second vaccination, respectively. With regard to underlying diseases, after the first vaccination, the lung cancer patients showed a significantly lower rates of seropositivity than non-cancer patients (38% vs 76%, p<0.001, on Architect; 45% vs 76%, p=0.005, on Elecsys). After the second vaccine dose, the percentage of positive seropositivity with non-cancer and lung cancer showed no difference between the two groups (100% vs 98%, p=1.000 on Architect; 100% and 98%, p=1.000 on Elecsys). However, the percentage of seroprotection in lung cancer patients was lower than that in non-cancer patients for the cutoff points of ≥ 1,084 AU/mL (64% vs 87%, p=0.017) on Architect and ≥ 150 U/mL (66% and 90%, p=0.013) on Elecsys. In addition, there were significant differences in the percentages of seroprotection after the second vaccination among the types of anticancer treatments. Notably, the percentages of seroprotection in lung cancer patients with cytotoxic agents were low (43% for ≥ 1,084 AU/mL, 0% for ≥ 5458 AU/mL, on Architect; 57% for ≥ 150 U/mL, 0% for ≥ 753 U/mL on Elecsys), whereas those in lung cancer patients receiving ICIs were high (82% for ≥ 1,084 AU/mL, 46% for ≥ 5458 AU/mL, on Architect; 82% for ≥ 150 U/mL, 46% for ≥ 753 U/mL on Elecsys), which were similar to those in non-cancer patients.

**Table 3.**
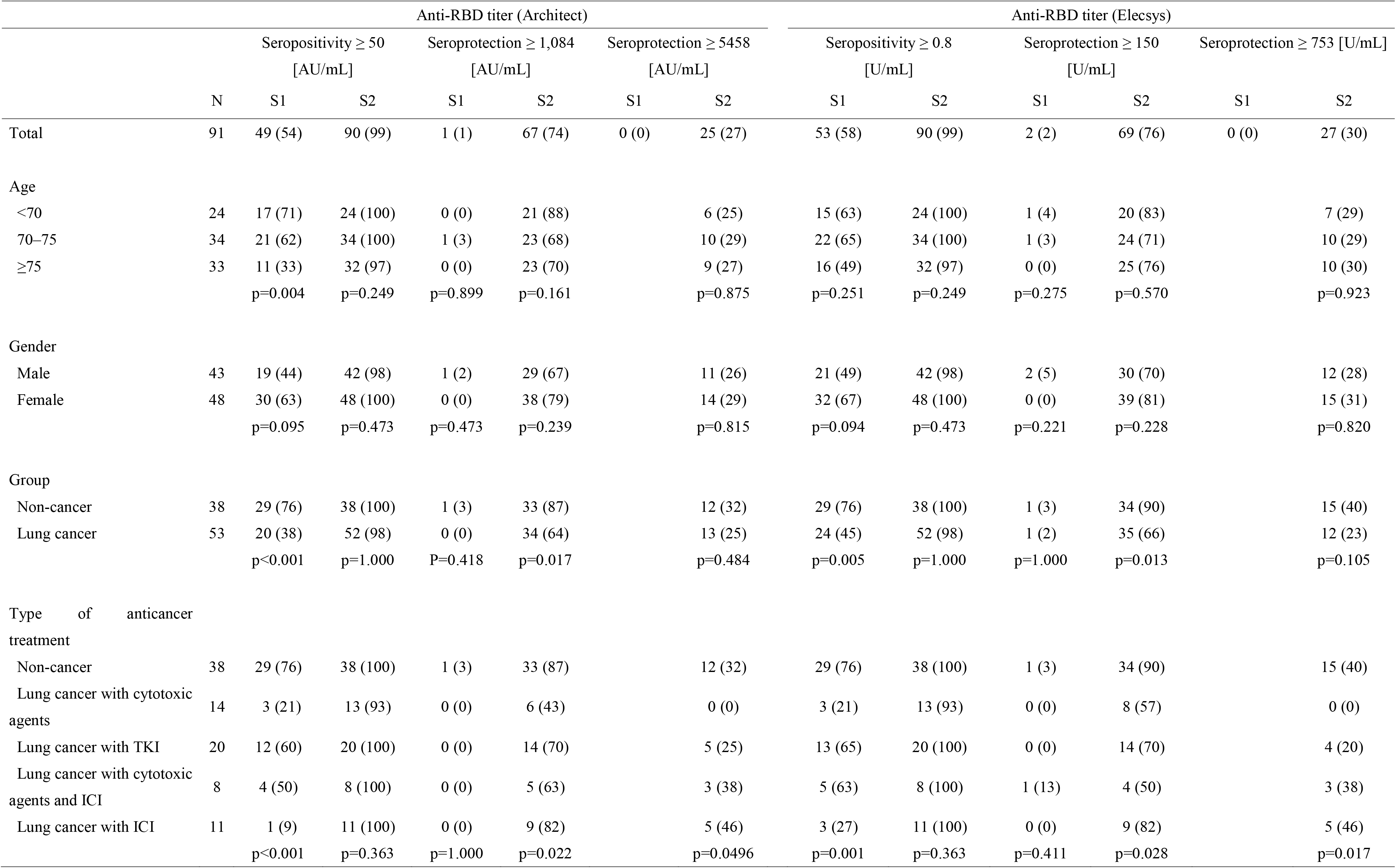
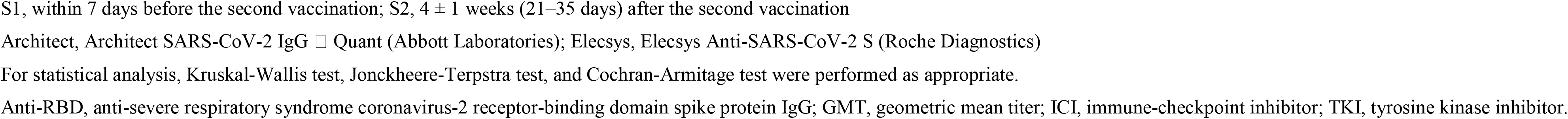
Seropositivity and seroprotection among study participants stratified by age, gender, group, and the type of anticancer treatment

Table 4 shows the adjusted ORs for seropositivity and seroprotection after vaccination with the BNT162b2 vaccine. The ORs for seropositivity in lung cancer patients were significantly lower than those in non-cancer patients (0.20, 95% CI, 0.08–0.55, on Architect; 0.29, 95% CI, 0.11–0.75, on Elecsys) after the first vaccination. The ORs for seroprotection was significant reduced in lung cancer patients after the second vaccination for the cutoff value of ≥ 1,084 AU/mL on Architect and ≥ 150 U/mL on Elecsys showing 0.25 (95% CI 0.08–0.78) and 0.21 (95% CI 0.06–0.71), respectively. Among the anticancer treatment types, the ORs for seropositivity in lung cancer patients receiving cytotoxic agents were reduced: 0.07 (95% CI: 0.01–0.37) for Architect and 0.09 (95% CI 0.02–0.44) for Elecsys after the first vaccination and 0.08 (95% CI: 0.01–0.41) for ≥ 1,084 AU/mL on Architect and 0.14 for ≥ 150 U/mL (95% CI 0.03–0.75) on Elecsys after the second vaccination. In contrast, the OR in lung cancer patients receiving ICIs after the second vaccination was 0.73 (0.11–4.99) for ≥ 1,084 U/mL on Architect and 0.54 (95% CI 0.07–3.83) for ≥ 150 U/mL on Elecsys, showing no significant difference compared with that in non-cancer patients.

**Table 4.**
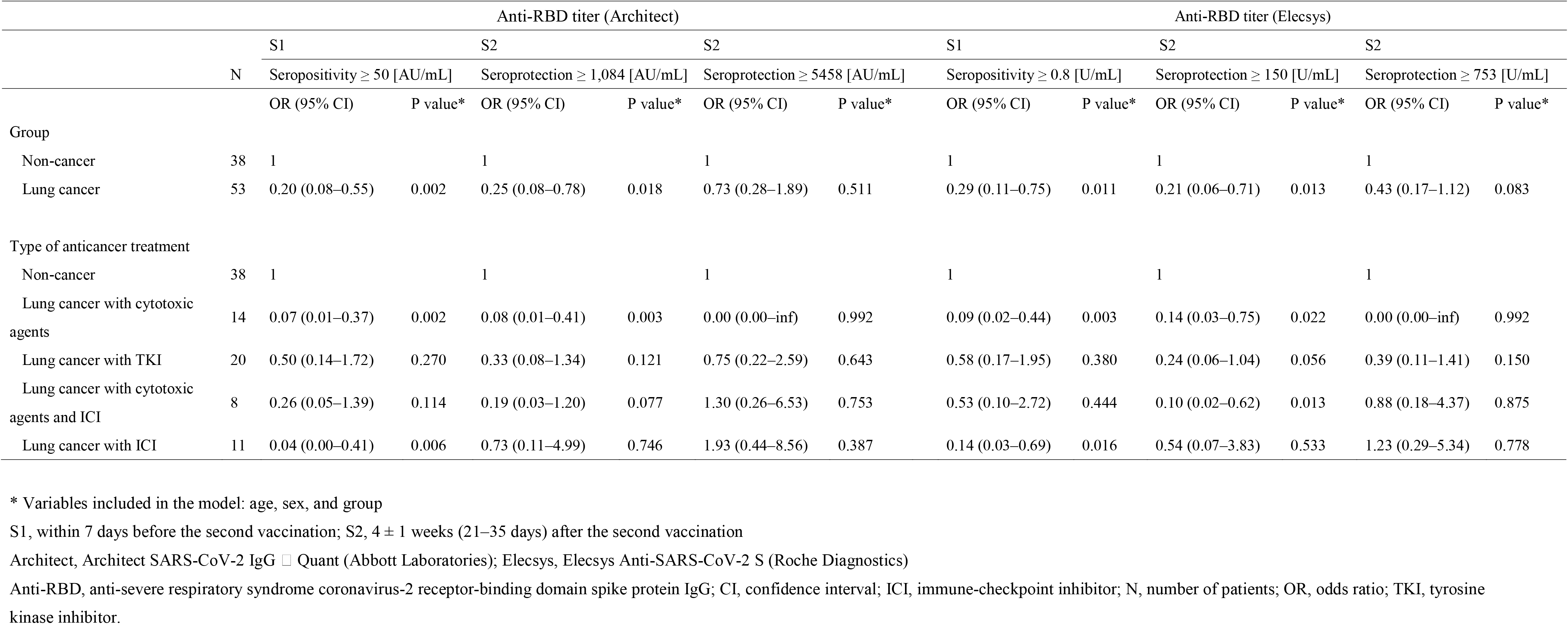
Adjusted odds ratios for seropositivity and seroprotection after COVID-19 vaccination in non-cancer patients and lung cancer patients

Table 5 shows the adverse reactions to the BNT162b2 COVID-19 vaccination. There were no significant differences in ocular or respiratory symptoms within 48 h, local reactions (within 48 h, and 48 h to 1 week), or systemic reactions (within 48 h, and 48 h to 1 week) between lung cancer patients and non-cancer patients after the first vaccination. Local reactions within 48 h after vaccination were observed in 79% of lung cancer patients and 81% of non-cancer patients (p=1.000). The most frequent local reaction within 48 h after the first vaccination was pain (72% in lung cancer patients and 78% in those with non-cancer patients, p=0.624). Forty-two percent of lung cancer patients and 47% of non-cancer patients had systemic reactions within 48 h after the first vaccination (p=0.666). No significant differences in adverse reactions existed after the second vaccination, except for itching being more common within 48 h in non-cancer patients. Local reactions within 48 h after the second vaccination increased overall compared with that after the first vaccination (83% in lung cancer patients, 84% in those with non-cancer patients, p=1.000). Systemic reactions within 48 h after the second vaccination also increased overall, compared with that after the first vaccination (65% of lung cancer patients, 68% of those with underlying disease, p=1.000). The frequencies of fever (27% vs. 19%, p=0.454), fatigue (36% vs. 43%, p=0.516), and myalgia (26% vs. 38%, p=0.259) in lung cancer patients vs. non-cancer patients, respectively, were particularly high. Most cases were mild and did not affect daily life.

**Table 5.** Adverse reactions to the BNT162b2 COVID-19 vaccination

|  | After 1<sup>st</sup> vaccination |  |  |  |  |  | After 2<sup>nd</sup> vaccination |  |  |  |  |  |
| --- | --- | --- | --- | --- | --- | --- | --- | --- | --- | --- | --- | --- |
|  | Within 48 h |  |  | 48 h–1 wk |  |  | Within 48 hours |  |  | 48 h–1 wk |  |  |
|  | Lung cancer | Non-cancer | P value | Lung cancer | Non-cancer | P value | Lung cancer | Non-cancer | P value | Lung cancer | Non-cancer | P value |
| Oculorespiratory syndrome |  |  |  |  |  |  |  |  |  |  |  |  |
| Total | 15 (8/53) | 14 (5/37) | 1.000 |  |  |  | 19 (10/53) | 19 (7/37) | 1.000 |  |  |  |
| Red eyes | 2 (1/53) | 3 (1/37) | 1.000 |  |  |  | 2 (1/53) | 3 (1/37) | 1.000 |  |  |  |
| Facial edema | 0 (0/53) | 0 (0/37) | 1.000 |  |  |  | 2 (1/53) | 3 (1/37) | 1.000 |  |  |  |
| Respiratory symptoms | 15 (8/53) | 16 (6/38) | 1.000 |  |  |  | 17 (9/53) | 21 (8/37) | 0.786 |  |  |  |
| Local reactions |  |  |  |  |  |  |  |  |  |  |  |  |
| Total | 79 (42/53) | 81 (30/37) | 1.000 | 21 (11/53) | 16 (6/37) | 0.785 | 83 (44/53) | 84 (31/37) | 1.000 | 43 (23/53) | 41 (15/37) | 0.831 |
| Redness | 8 (4/53) | 8 (3/37) | 1.000 | 2 (1/53) | 3 (1/37) | 1.000 | 9 (5/53) | 16 (6/37) | 0.349 | 9 (5/53) | 19 (3/37) | 0.220 |
| Swelling | 15 (8/53) | 19 (7/37) | 0.775 | 6 (3/53) | 5 (2/37) | 1.000 | 21 (11/53) | 32 (12/37) | 0.229 | 19 (10/53) | 16 (6/37) | 0.787 |
| Induration | 8 (4/53) | 22 (8/37) | 0.065 | 4 (2/53) | 3 (1/37) | 1.000 | 11 (6/53) | 27 (10/37) | 0.091 | 9 (5/53) | 16 (6/37) | 0.349 |
| Pain | 72 (38/53) | 78 (29/37) | 0.624 | 19 (10/53) | 14 (5/37) | 0.576 | 78 (41/53) | 81 (30/37) | 0.795 | 28 (15/53) | 32 (12/37) | 0.816 |
| Itch | 4 (2/53) | 0 (0/37) | 0.510 | 4 (2/53) | 5 (2/37) | 1.000 | 8 (4/53) | 24 (9/37) | 0.034 | 11 (6/53) | 16 (6/37) | 0.541 |
| Heat | 26 (14/53) | 27 (10/37) | 1.000 | 8 (4/53) | 3 (1/37) | 0.645 | 23 (12/53) | 43 (16/37) | 0.063 | 15 (8/53) | 19 (7/37) | 0.775 |
| Systemic reactions |  |  |  |  |  |  |  |  |  |  |  |  |
| Total | 42 (22/53) | 47 (17/36) | 0.666 | 25 (13/53) | 9 (3/34) | 0.090 | 65 (34/52) | 68 (25/37) | 1.000 | 35 (18/51) | 36 (13/36) | 1.000 |
| Fever | 15 (8/53) | 14 (5/36) | 1.000 | 6 (3/53) | 0 (0/34) | 0.277 | 27 (14/52) | 19 (7/37) | 0.453 | 14 (7/51) | 9 (3/37) | 0.510 |
| Fatigue | 19 (10/53) | 19 (7/37) | 1.000 | 11 (6/53) | 8 (3/37) | 0.732 | 36 (19/53) | 43 (16/37) | 0.516 | 21 (11/53) | 24 (9/37) | 0.798 |
| Myalgia | 25 (13/53) | 32 (12/37) | 0.477 | 8 (4/53) | 8 (3/37) | 1.000 | 26 (14/53) | 38 (14/37) | 0.259 | 8 (4/53) | 16 (6/37) | 0.307 |
| Joint pain | 8 (4/53) | 8 (3/37) | 1.000 | 4 (2/53) | 0 (0/37) | 0.510 | 15 (8/53) | 8 (3/37) | 0.515 | 4 (2/53) | 8 (3/37) | 0.398 |
| Headache | 4 (2/53) | 14 (5/37) | 0.119 | 4 (2/53) | 8 (3/37) | 0.398 | 13 (7/53) | 16 (6/37) | 0.765 | 4 (2/53) | 8 (3/37) | 0.398 |
| Chills | 2 (1/53) | 0 (0/37) | 1.000 | 0 (0/53) | 0 (0/37) | 1.000 | 15 (8/53) | 5 (2/37) | 0.188 | 2 (1/53) | 3 (1/37) | 1.000 |
| Nausea | 2 (1/53) | 0 (0/37) | 1.000 | 0 (0/53) | 0 (0/37) | 1.000 | 4 (2/53) | 0 (0/37) | 0.510 | 0 (0/53) | 0 (0/37) | 1.000 |
| Diarrhea | 2 (1/53) | 5 (2/37) | 0.566 | 4 (2/53) | 0 (0/37) | 0.510 | 8 (4/53) | 5 (2/37) | 1.000 | 9 (5/53) | 0 (0/37) | 0.075 |
| Rash | 2 (1/53) | 0 (0/37) | 1.000 | 2 (1/53) | 0 (0/37) | 1.000 | 4 (2/53) | 3 (1/37) | 1.000 | 6 (3/53) | 3 (1/37) | 0.641 |

Note: Data are presented as percent (n/N).
For the calculation of P value, Fisher's exact test was performed

## Discussion

We evaluated the immunogenicity and safety of the BNT162b2 COVID-19 vaccine after the first and second vaccinations in lung cancer patients and compared them with those in non-cancer patients. Lung cancer patients showed a significant increase in the GMT; however, the GMT was significantly lower in these patients than in non-cancer patients. In the multivariate analysis, the adjusted OR for seropositivity and seroprotection (_≥_1,084 AU/mL for Architect and _≥_ 150 AU/mL for Elecsys) by the BNT162b2 vaccine was significantly lower in lung cancer patients than in non-cancer patients. In the analysis of the anticancer treatment types, the adjusted OR for seropositivity and seroprotection (_≥_ AU/mL for Architect and _≥_ 150 AU/mL for Elecsys) was significantly lower in lung cancer patients receiving cytotoxic agents than in non-cancer patients. Additionally, there was no increase in the number of adverse reactions in lung cancer patients compared with that in non-cancer patients.

Several studies have shown that the immunogenicity of the COVID-19 vaccine is reduced in cancer patients; moreover, inadequate antibody responses have been reported, especially in cancer patients vaccinated with a single dose.^12, 21, 22^ In a meta-analysis, compared with controls, cancer patients with incomplete vaccination showed a 55% reduced likelihood of being seropositive for anti-spike IgG titers, whereas those completely vaccinated had a 31% reduced likelihood of seroconversion.^21^ A lower seroconversion rate of anti-spike IgG antibody was reported especially in patients with hematologic malignancies and those receiving the anti-CD20 antibody.^21, 23, 24^ Conversely, patients with solid tumors may have a sufficient seroconversion rate after two doses of mRNA vaccination. In a meta-analysis, 94% of patients with solid tumors demonstrated positive antibodies after two vaccination doses.^21^ Studies using the same Abbott reagent for anti-RBD antibodies, as in our study, reported that completely vaccinated patients with solid tumors showed 90–98% of seropositivity at a cutoff value ≥ 50 AU/mL.^12, 14, 23^ Here, we found 98% of seropositivity at a cutoff value ≥ 50 AU/mL in lung cancer patients receiving anticancer treatment. Nevertheless, patients with solid tumors reportedly had a lower titer of anti-spike IgG than healthy subjects, even after complete vaccination.^12, 14, 25^ Here, the GMT was significantly lower in lung cancer patients than in non-cancer patients, considering a stratified analysis.

Recent studies have shown an association between antibody titer and vaccine efficacy.^17-19, 26, 27^ In this study, we adopted two cutoff points for seroprotection according to previous studies.^17, 19^ The cutoff value of ≥ 154 BAU/mL seemed reasonable for the analysis of immunocompromised patients, such as cancer patients, because of the fact that the calculation of ORs in lung cancer patients receiving cytotoxic agents was impossible owing to no seroprotection for ≥ 775 BAU/mL (converted to 5458 AU/mL for Architect and 753 U/mL for Elecsys) in this study. There was no difference in the percentage of seropositivity between lung cancer patients and non-cancer patients for a cutoff value ≥ 50 AU/mL after two doses of vaccination; however, the seroprotection in lung cancer patients was lower than that in non-cancer patients with cutoff values of ≥ 1,084 AU/mL and ≥ 150 AU/mL for Architect and Elecsys, respectively. Furthermore, a significant decrease was observed in the adjusted OR for seroprotection with cutoff values of ≥ 1,084 AU/mL and ≥ 150 AU/mL for Architect and Elecsys, respectively. Compared to that in non-cancer patients, the immunogenicity of the COVID-19 vaccine in lung cancer patients undergoing anticancer treatment could be inadequate.

Several studies reported that cytotoxic agents reduce the immunogenicity of COVID-19 vaccines in patients with solid cancer.^14, 25, 28-30^ Consistent with the results of prior studies, in this study, the OR for seroprotection at cutoff values of ≥ 1,084 AU/mL for Architect and ≥ 150 U/mL for Elecsys after two vaccination doses was significantly decreased in lung cancer patients undergoing treatment with cytotoxic agents. Additionally, there are limited data on whether TKIs affect the immunogenicity of COVID-19 vaccines.^14, 28^ In a study evaluating patients with thoracic cancer using the same Abbott reagent used in the present study, treatment with TKIs was associated with reduced antibody response to BNT162b2 COVID-19 vaccine compared with that in health controls.^14^ Similarly, in our study, the adjusted OR for ≥ 1,084 AU/mL on Architect and ≥ 150 AU/mL on Elecsys after two vaccination doses tended to decrease in patients treated with TKIs, although the result was statistically nonsignificant. Several studies have reported that ICIs do not decrease the immunogenicity of COVID-19 vaccines.^23, 31^ Notably, the adjusted ORs for seroprotection in patients receiving ICIs were 0.73 (0.11–4.99) for ≥ 1,084 U/mL on Architect and 0.54 (95% CI 0.07–3.83) for ≥ 150 U/mL on Elecsys after the second vaccination, which did not decrease as compared with that in non-cancer patients.

Interestingly, the GMT ratio of S2/S0 was significantly different between groups and among types of anticancer treatment on Elecsys whereas it was insignificant on Architect, showing reduced immunogenicity on Elecsys compared with that on Architect. This discrepancies in results between the two assays might be because of the different methods used to identify the antibodies.^32^ First, the repertoire of the antibodies might affects the titers measured in each immunoassay ^33^. Second, Elecsys uses the double-antigen sandwich method, whereas Architect uses a chemiluminescent microparicle immunoassay^15, 16^ The Elecsys platform based on the double-antigen sandwich method has shown tendency towards detecting antibodies with higher avidity.^34^ Thus, the accentuated intergroup difference in GMT ratio observed on the Elecsys platform may be a reflection of a potential defect in the antibodies’ affinity maturation process in patients with lung cancer. High avidity IgG has been reported to play an important role in immunity against SARS-CoV-2.^35^ Further studies are needed to evaluate the kinetics of antibody avidity in immunocompromised patients, such as cancer patients, after vaccination.

The short-term safety of the COVID-19 vaccine in cancer patients undergoing treatment is reportedly comparable to that in healthy subjects.^25^ In a review of seven studies—three including patients with hematologic malignancies, two including those with solid tumors, and two including those with both types of cancer—the authors reported that the frequency of local and systemic reactions did not differ between cancer patients and healthy subjects after partial and full vaccination.^25^ A large cohort study that evaluated 1753 patients, including 1094 with solid tumors and 89 with hematologic malignancies, revealed few differences in the frequency of adverse reactions between cancer patients and non-cancer patients.^36^ Here, no increase in adverse reactions was observed in patients with lung cancer after either the first or second vaccination dose.

Here, we observed a significant increase in antibody titers and an acceptable safety level of BNT162b2 COVID-19 vaccination in lung cancer patients undergoing anticancer treatment, which could support the use of COVID-19 vaccination in this patient group. However, compared with that in non-cancer patients, immunogenicity may be inadequate; moreover, further studies are needed to determine whether increased doses, the vaccine type, mixing vaccine types, the timing of vaccination, or additional doses enhance immunogenicity in cancer patients under treatment. Cancer patients should continue to be prioritized for vaccination. Studies evaluating the immunogenicity of COVID-19 vaccination in cancer patients are increasing in number; however, few have evaluated clinical outcomes such as the incidence of infection, hospitalization, and death.^21, 37, 38^ Further studies are needed to evaluate the effectiveness of COVID-19 vaccination in cancer patients considering the various cancer types, administered vaccine, type of anticancer treatment, number of doses, timing of vaccination, mixed vaccination, immunogenicity by assay method, and clinical outcomes such as incidence of COVID-19 and death. Infection control measures, including universal mask wearing and social distancing, continue to be important for lung cancer patients undergoing anticancer treatment.

Our study had some limitations. First, the sample size was small. Nevertheless, this was a two-center multicenter study, and only few studies have evaluated only lung cancer patients. The sample size of 53 lung cancer patients was relatively large as compared with those in previous studies, and we were able to undertake a multivariate analysis considering various types of anticancer treatment. Second, we did not evaluate the cellular immunity associated with the COVID-19 vaccination. However, the anti-RBD antibody titer evaluated here is reportedly correlated with the efficacy of the COVID-19 vaccination.^17, 18, 26^ Third, only short-term immunogenicity and safety were evaluated here. However, this study is ongoing, and antibody titers and adverse events, including immune-related events, 6 months after the second vaccination will be reported in a future paper. Further studies and infection control measures should be considered to protect patients with lung cancer from COVID-19.

## Supporting information

CRediT Statement

## Data Availability

All data produced in the present study are available upon reasonable request to the authors

## Abbreviations

CI,: confidence interval;
COVID-19,: coronavirus disease;
GMT,: geometric mean antibody titer;
ICIs,: immune-checkpoint inhibitors;
OR,: odds ratio;
sP,: seroprotection rate;
TKIs;: tyrosine kinase inhibitors.

## Acknowledgments

We thank all the staff of the Department of Pulmonary Medicine and Laboratory Medicine at Kameda Medical Center and Chikamori Hospital for their involvement in this study.

