## Supplementary Figure 1 for "Immunogenicity and safety of COVID-19 vaccine in lung cancer patients receiving anticancer treatment: A prospective multicenter cohort study"

Supplemental Figure 1 Changes in the anti-RBD antibody titer before vaccination (S0), after first vaccination (S1), and after second vaccination (S2) among types of anticancer treatment. Reduced anti-RBD titers in lung cancer patients receiving cytotoxic agents after the second vaccination (S2) is more pronounced in the results of Elecsys assay.

Anti-RBD, anti-severe respiratory syndrome coronavirus-2 receptor-binding domain spike protein IgG; LC, lung cancer; ICI, immune-checkpoint inhibitor; TKI, tyrosine kinase inhibitor

S0, within 14 days before the first vaccination; S1, within 7 days before the second vaccination; S2, 4 ± 1 weeks (21–35 days) after the second vaccination

Architect, Architect SARS-CoV-2 IgG Ⅱ Quant (Abbott Laboratories); Elecsys, Elecsys Anti-SARS-CoV-2 S (Roche Diagnostics)

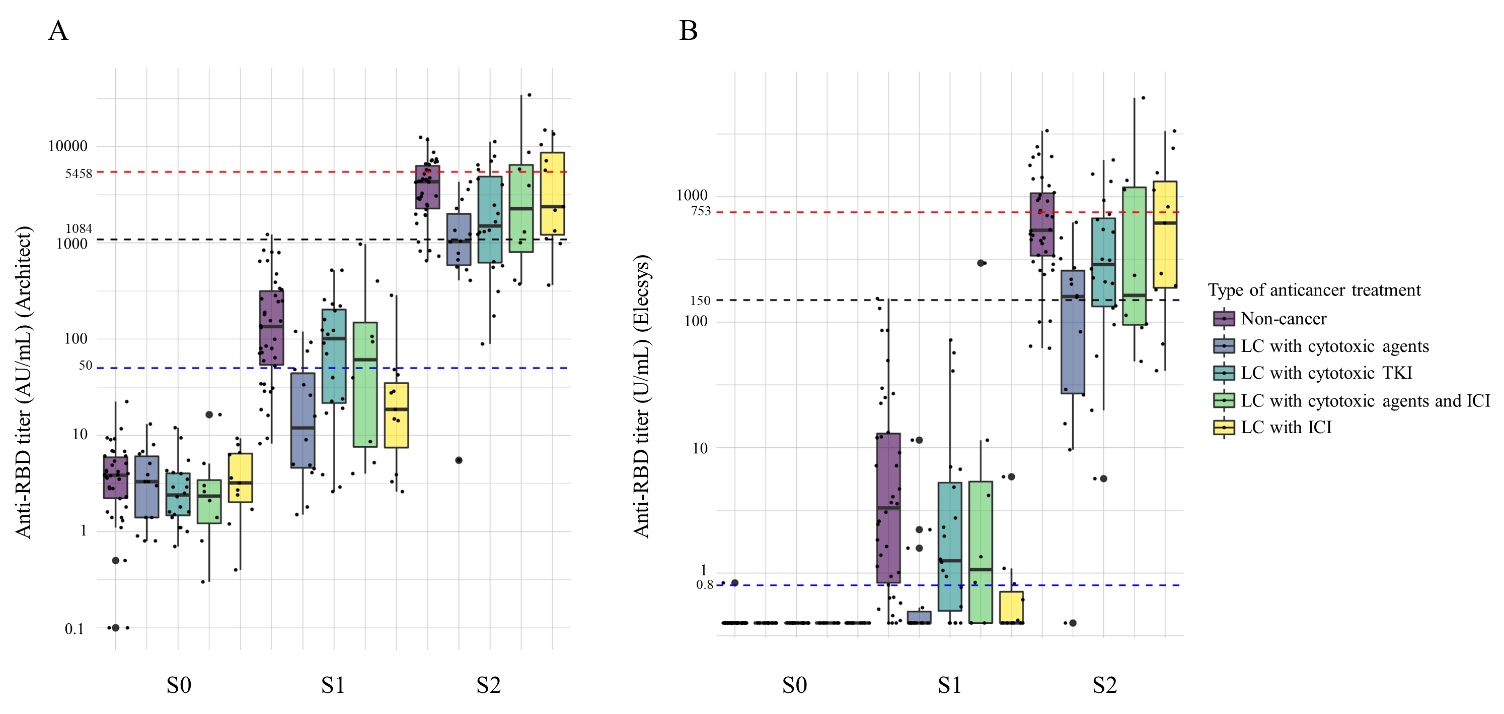
