## Supplementary material for "Immunogenicity and safety of COVID-19 vaccine in lung cancer patients receiving anticancer treatment: A prospective multicenter cohort study": CRediT Statement

K.N. contributed to conceptualization, data curation, formal analysis, investigation, methodology, project administration, software, validation, visualization, writing – original draft, and writing – review & editing. M.I. contributed to conceptualization, investigation, resources, methodology, project administration, and writing – review & editing. H.M. contributed to conceptualization, data curation, formal analysis, methodology, software, validation, visualization, and writing – review & editing. C.Y., T.N. M.S., H.N., and Y.O contributed to investigation, resources, and writing – review & editing. Y.N. contributed to funding acquisition, investigation, resources, visualization, and writing – review & editing. N.K. and Y.N. contributed to investigation, resources, and writing – review & editing. Y.K. contributed conceptualization, funding acquisition, investigation, resources, visualization, and writing – review & editing. Y.H. contributed to conceptualization, funding acquisition, methodology, supervision, and writing – review & editing. All authors have read and approved the final draft of the article.
